# Novel CSF biomarkers of GLUT1 deficiency syndrome: implications beyond the brain’s energy deficit

**DOI:** 10.1101/2022.04.15.22273511

**Authors:** Tessa M.A. Peters, Jona Merx, Pieter C. Kooijman, Marek Noga, Siebolt de Boer, Loes A. van Gemert, Guido Salden, Udo F.H. Engelke, Dirk J. Lefeber, Rianne E. van Outersterp, Giel Berden, Thomas J. Boltje, Rafael Artuch, Leticia Pías, Ángeles García-Cazorla, Ivo Barić, Beat Thöny, Jos Oomens, Jonathan Martens, Ron A. Wevers, Marcel M. Verbeek, Karlien L.M. Coene, Michèl A.A.P. Willemsen

## Abstract

We used next-generation metabolic screening to identify new biomarkers for improved diagnosis and pathophysiological understanding of glucose transporter type 1 deficiency syndrome (GLUT1DS), comparing metabolic CSF profiles from 11 patients to those of 116 controls. This confirmed decreased CSF glucose and lactate levels in patients with GLUT1DS and increased glutamine at group level. We identified three novel biomarkers significantly decreased in patients, namely gluconic + galactonic acid, xylose-α1-3-glucose and xylose-α1-3-xylose-α1-3-glucose, of which the latter two have not previously been identified in body fluids. CSF concentrations of gluconic + galactonic acid may be reduced as these metabolites could serve as alternative substrates for the pentose phosphate pathway. Xylose-α1-3-glucose and xylose-α1-3-xylose-α1-3-glucose may originate from *O-*glycosylated proteins; their decreased levels are hypothetically the consequence of insufficient glucose, one of two substrates for *O*-glucosylation. Since many proteins are *O*-glucosylated, this deficiency may affect cellular processes and thus contribute to GLUT1DS pathophysiology. The novel CSF biomarkers have the potential to improve the biochemical diagnosis of GLUT1DS. Our findings imply that brain glucose deficiency in GLUT1DS may cause disruptions at the cellular level that go beyond energy metabolism, underlining the importance of developing treatment strategies that directly target cerebral glucose uptake.

## Introduction

Glucose transporter type 1 deficiency syndrome (GLUT1DS) is an inborn error of metabolism (IEM), first documented in 1991.^1^ As glucose transport into the brain depends on GLUT1,^2^ a defect of this transporter results in chronic neuroglycopenia. Classically, GLUT1DS is characterized by an infantile onset encephalopathy with intellectual disability, movement disorders, and drug-resistant epilepsy.^1,3,4^ Nowadays, many patients have been recognized who suffer from only one or two of these core features, sometimes with onset as late as in adulthood.^3-5^ The transporter defect leads to low CSF concentrations of glucose and lactate, in the context of normoglycaemia.^1,3-5^ The first-line treatment of GLUT1DS is ketogenic diet therapy (KDT), which provides the brain with alternative energy sources.^4^ For unknown reasons, however, not all patients respond to KDT,^6^ and existing cognitive defects generally remain present despite adequate seizure control.

The *SLC2A1* gene encodes GLUT1, and heterozygous variants of this gene can be demonstrated in most patients.^3,4^ Of note, CSF glucose and lactate and the CSF/blood glucose ratio can be used for diagnosis, but may be close to or within reference ranges, especially in mild cases.^3-5,7^ Since erythrocytes also use GLUT1 as the major glucose transporter, techniques that quantify the amount of GLUT1 protein on the erythrocyte surface^8^ or measure its transmembrane transport capacity^9^ may be of additional diagnostic use. These techniques, however, face pre-analytical challenges and do not provide information about the consequences of the defect for the brain.

Next-generation metabolic screening (NGMS) is a reversed-phase liquid chromatography-mass spectrometry (RPLC-MS)-based untargeted metabolomics approach, which is used for diagnostic screening of IEMs.^10,11^ We applied NGMS to CSF of GLUT1DS patients in order to investigate its potential as a functional test for this diagnosis, with the aim to increase diagnostic accuracy. The untargeted nature of NGMS offers the possibility to expand the GLUT1DS CSF profile beyond glucose and lactate, thereby increasing our biochemical understanding of the underlying disease mechanisms and guiding therapy development.

## Materials and methods

### Sample selection

CSF samples from patients with GLUT1DS were retrospectively included when at least two of the three following criteria were met: (1) classical phenotype, (2) typical CSF profile of GLUT1DS,^5^ and (3) a pathogenic *SLC2A1* variant. This resulted in a selection of samples from 11 patients, aged 1 to 20 years at time of sample collection (Table 1). For controls, we used the CSF samples from our previously described control cohort (aged 0-15 years; *n* = 87),^11^ extended with 29 samples from patients with other neurometabolic diseases. First, we studied a ‘discovery cohort’ of eight patients with GLUT1DS (Patients 1-8) and 15 (out of the 116) age-matched controls in one single experiment. At later timepoints, CSF samples from the ‘validation cohort’ of three additional GLUT1DS patients (Patients 9-11) and the remaining controls were measured in four different runs.

**Table 1.** Patient characteristics and diagnostic value of selected features in CSF.

| Pt | Sex | Age group<sup>a</sup> | Variant in *SLC2A1* gene | CSF glucose (ref) (mM)<sup>b</sup> | CSF lactate (ref) (μM)<sup>b</sup> | CSF/blood glucose ratio (ref)<sup>b</sup> | Clinical phenotype<sup>a</sup> | Treatment<sup>a</sup> | Normalized intensity GluA+GalA (ref: 0.268-1.241)<sup>c</sup> | Normalized intensity Xyl-Glc (ref: 0.459-1.890)<sup>c</sup> | Normalized intensity Xyl-Xyl-Glc (ref: 0.279-2.705)<sup>c</sup> |
| --- | --- | --- | --- | --- | --- | --- | --- | --- | --- | --- | --- |
| 1 | F | 6-10 | c.457C>T | **2.0 (2.6-3.8)** | **1090 (1170-1850)** | **0.37 (0.49-0.79)** | Classical<sup>d</sup> | None | 0.305 | **0.295** | **0.141** |
| 2 | F | 0-5 | c.493G>A | **1.9 (2.6-3.8)** | **968 (1090-1800)** | **0.37 (0.49-0.79)** | Classical<sup>d</sup> | None | 0.293 | **0.354** | 0.466 |
| 3 | F | 16-20 | c.632C>A | **2.1 (2.2-3.4)** | **890 (1000-2200)** | **0.39 (na)<sup>e</sup>** | Classical<sup>d</sup> | None | **0.266** | 0.584 | 0.584 |
| 4 | F | 16-20 | c.632C>A | 2.4 (2.2-3.4) | **770 (1000-2200)** | **0.46 (na)<sup>e</sup>** | Classical<sup>d</sup> | None | **0.201** | **0.320** | **0.116** |
| 5 | F | 10-15 | c.524G>T | **1.8 (2.2-3.4)** | **810 (1000-2200)** | na | Unknown | Unknown | **0.238** | **0.248** | **0.161** |
| 6 | M | 10-15 | c.998G>A | 2.2 (2.2-3.4) | 1100 (1000-2200) | **0.42 (na)<sup>e</sup>** | Classical<sup>d</sup> | None | 0.272 | **0.408** | **0.099** |
| 7 | M | 0-5 | c.277C>T | **1.5 (2.2-3.4)** | **800 (1000-2200)** | **0.40 (na)<sup>e</sup>** | Classical<sup>d</sup> | None | **0.198** | **0.206** | **0.164** |
| 8 | M | 6-10 | Missense (na) | **1.1 (2.6-3.8)** | **541 (1170-1850)** | na | Classical<sup>d</sup> | None | **0.178** | 1.007 | **0.257** |
| 9 | M | 6-10 | c.940G>A | **2.4 (2.6-3.8)** | 1210 (1170-1850) | 0.50 (0.49-0.79) | Classical<sup>d</sup> | None | **0.243** | **0.214** | 0.283 |
| 10 | F | 6-10 | c.1390G>C | **2.3 (2.7-3.9)** | 1310 (1260-1990) | **0.42 (0.50-0.81)** | Classical<sup>d</sup> | Ketogenic diet | **0.164** | **0.252** | 0.340 |
| 11 | M | 6-10 | c.998G>A | **2.0 (2.7-4.0)** | 1830 (1200-2100) | **0.29 (>0.6)** | Classical<sup>d</sup> | Valproic acid | **0.207** | **0.189** | **0.117** |

Boldfaced values are outside reference range.
<sup>a</sup>Age group (in years), phenotype, and treatment at moment of lumbar puncture.
<sup>b</sup>Reference ranges given between brackets are provided by the laboratory that performed the CSF analysis.
<sup>c</sup>Reference range is defined by p5-p95 of the control population.
<sup>d</sup>Combination of mild to moderate intellectual disability, epilepsy, and movement disorder.
<sup>e</sup>While no reference values are provided, this value is below the 10<sup>th</sup> percentile described by Leen et al. 2013,<sup>6</sup> fitting with GLUT1DS.
F = female, M = male, ref = reference range, na = not available, GluA/GalA = gluconic acid and galactonic acid, Xyl-Glc = xylose-α1,3-glucose, and Xyl-Xyl-Glc = xylose-α1,3-xylose-α1,3-glucose

The study was conducted in accordance with the Declaration of Helsinki. All patients (or their guardians) approved of the use of their left-over samples for method validation purposes (Radboud University Medical Center, ethical committee file 2016-3011).

### Chemical standards

The synthesis of diketogulonic acid (DKG), xylose-α1-3-glucose (Xyl-Glc) and xylose-α1-3-xylose-α1-3-glucose (Xyl-Xyl-Glc) is included in the Supplementary Material. All other chemical standards were purchased from Merck KGaA (Darmstadt, Germany).

### NGMS – targeted analysis

Samples were processed and measured in duplicate as previously described.^12,13^ Subsequently, we performed targeted data analysis for glucose and lactate, as well as other metabolites that have previously been reported as possible CSF GLUT1DS biomarkers: fructose, mannose, glutamine, inosine, glycerol-3-phosphate, and isocitrate.^12^ Furthermore, we included ascorbic acid, dehydroascorbic acid (DHA), and DKG,^13^ and galactose, glucosamine, 2-deoxy-D-glucose (2DOG), and 3-*O*-methyl-D-glucose (3OMG),^14^ as known substrates of GLUT1. For all these biomarkers, as well as xylose (see Discussion), we measured a reference standard, so that retention time (RT) could be used together with the accurate mass for “level 1” metabolite identification.^15^ Based on the molecular formula and RT, the raw RPLC-MS data were searched for these metabolites using Skyline (version 20.2.0.343). The corresponding intensities were extracted from all samples and further analysed using R version 4.1.1. Intensities of duplicate measurements were averaged and then normalized to the mean quality control (QC) intensity of the respective run. The fold change (FC) was calculated by dividing the median intensity in the patients by the median intensity in the controls. The means were compared using a Wilcoxon signed-rank test with Benjamini-Hochberg correction and were considered significantly different when the adjusted *P*-value (*q*) was below 0.05.

### NGMS – untargeted analysis

Raw data acquired from the RPLC-MS runs, from positive and negative ionization mode, were aligned using the R package “xcms” (XCMS version 3.4.4 running under R version 3.5.3). The extracted features (i.e. the combination of an accurate mass-to-charge-ratio (*m/z*), (RT), and intensity) were pre-processed by selecting those with *m/z* 70-700 and RT 0.4-16 min, with a procedure blank intensity lower than 10% of the mean intensity in the samples, and with an intensity of ≥ 50,000 counts in at least one sample.

After pre-processing, we compared feature intensities between patients and controls within the ‘discovery cohort’ using the Wilcoxon signed-rank test with Benjamini-Hochberg correction (significant at *q* < 0.05). Resulting significant features were excluded if (1) the corresponding raw data did not show a reliable peak, (2) they originated from known biomarkers (see sections on targeted analysis), (3) they were redundant because they represented the same metabolite as another selected feature (the feature with the highest non-saturated intensity was kept), or (4) they could not be reproduced in the ‘validation cohort’ (Patients 9-11). For the remaining features, intensities were extracted from the raw data, which were further analysed as described for targeted analysis.

For annotation of the selected features, we first compared the observed *m/z* and RT values to those present in our in-house IEM panel.^10^ If no matches were found, we searched the Human Metabolome Database (HMDB)^16^ based on *m/z* for the adducts [M+H]^+^ and [M+Na]^+^ (positive mode) and [M-H]^-^ and [M+Cl]^-^ (negative mode). Furthermore, we used Agilent MassHunter Qualitative Analysis 7.0.0 to predict a fitting molecular formula based on mass, isotope distance and relative isotope intensities, using the same adducts. Based on the putative annotations, ion-pair liquid chromatography tandem mass spectrometry (IP-LC-MS/MS) and hydrophilic interaction liquid chromatography infrared ion spectroscopy (HILIC-IRIS)^17^ were applied to confirm the identity of the metabolites. More details on these methods are desribed in the Supplementary Material.

To determine the diagnostic value of selected features, for each decreased or increased selected feature, the 5^th^ (95^th^) percentile (p5/p95) and 2.5^th^ (97.5^th^) percentile (p2.5/p97.5) were calculated based on the normalized intensities of the full control group.

### Data availability

The data that support the findings of this study are available from the corresponding author, upon reasonable request.

## Results

### Targeted analysis

For glycerol-3-phosphate, DHA, DKG, glucosamine and 2DOG, no matching features were found. Fructose, mannose and galactose are isobaric with glucose and have highly similar RPLC RTs. Therefore, the corresponding peaks were disrupted by the high signal of glucose and not useful for semiquantitative analysis. Results for glucose, lactate, glutamine, inosine, isocitric acid, ascorbic acid, 3OMG, and xylose are shown in Fig. 1. Glucose and lactate were significantly decreased in patients, while glutamine was significantly increased. The fold changes were -1.3, -1.2 and +1.4, respectively. For the remaining metabolites, no differences were found. Of note, ascorbic acid intensities were near the detection limit of the assay in many samples, with a high variation, probably due to limited stability of this metabolite.

**Figure 1.**
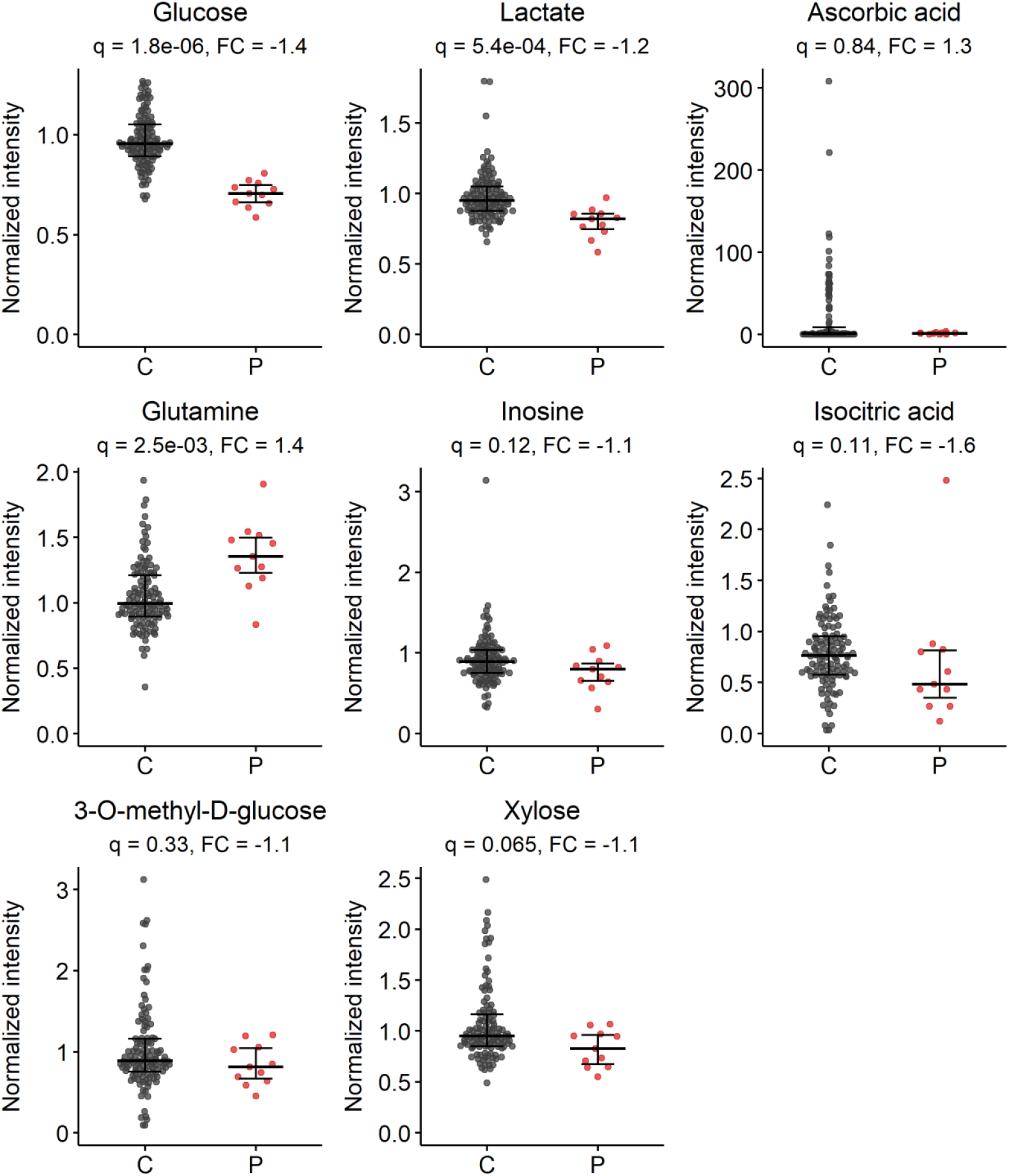
Normalized intensities of metabolites from targeted analysis. Metabolites include glucose (^13^C isotope of [M+Cl]^-^ adduct, *m/z* 216.0362), lactate (^13^C isotope of the [M-H]^-^ adduct, *m/z* 90.0278), ascorbic acid ([M+H]^+^ adduct, *m/z* 177.0394), glutamine (^13^C [M-H]^-^ adduct, *m/z* 145.0619), inosine ([M+H]^+^ adduct, *m/z* 269.0880), isocitric acid ([M+Na]^+^ adduct, *m/z* 215.0162), 3-*O*-methyl-D-glucose ([M+Na]^+^ adduct, *m/z* 217.0683), and xylose ([M+Cl]^-^ adduct, *m/z* 185.0222). Data include all control (C) and patient (P) samples (n = 116 and n = 11, respectively). Bars represent median with interquartile range. *q* = adjusted *P*-value (Wilcoxon signed-rank test with Benjamini-Hochberg correction), FC = fold change (calculated by median(P)/median(C)).

### Untargeted analysis

Using the data of the ‘discovery cohort’, feature extraction and alignment by XCMS yielded 33,135 features. After pre-processing, 7,618 features remained, of which 19 were significantly different between patients and controls. Subsequently, we excluded 16 features that originated from glucose or lactate, could not be reproduced in the raw data, were redundant, or were not reproducible across all measurements. Three remaining features (n1905, n5938 and p16243, with n = negative and p = positive MS-mode) were selected for further analysis. These all showed decreased intensities in patients compared to controls. Using the intensities from the raw data, the significant difference between patients and controls in the ‘discovery cohort’ was confirmed for all three features (*q* < 0.0001; Fig. 2). Moreover, these findings could be reproduced in the independent ‘validation cohort’ (*q* < 0.001; Fig. 2).

**Figure 2.**
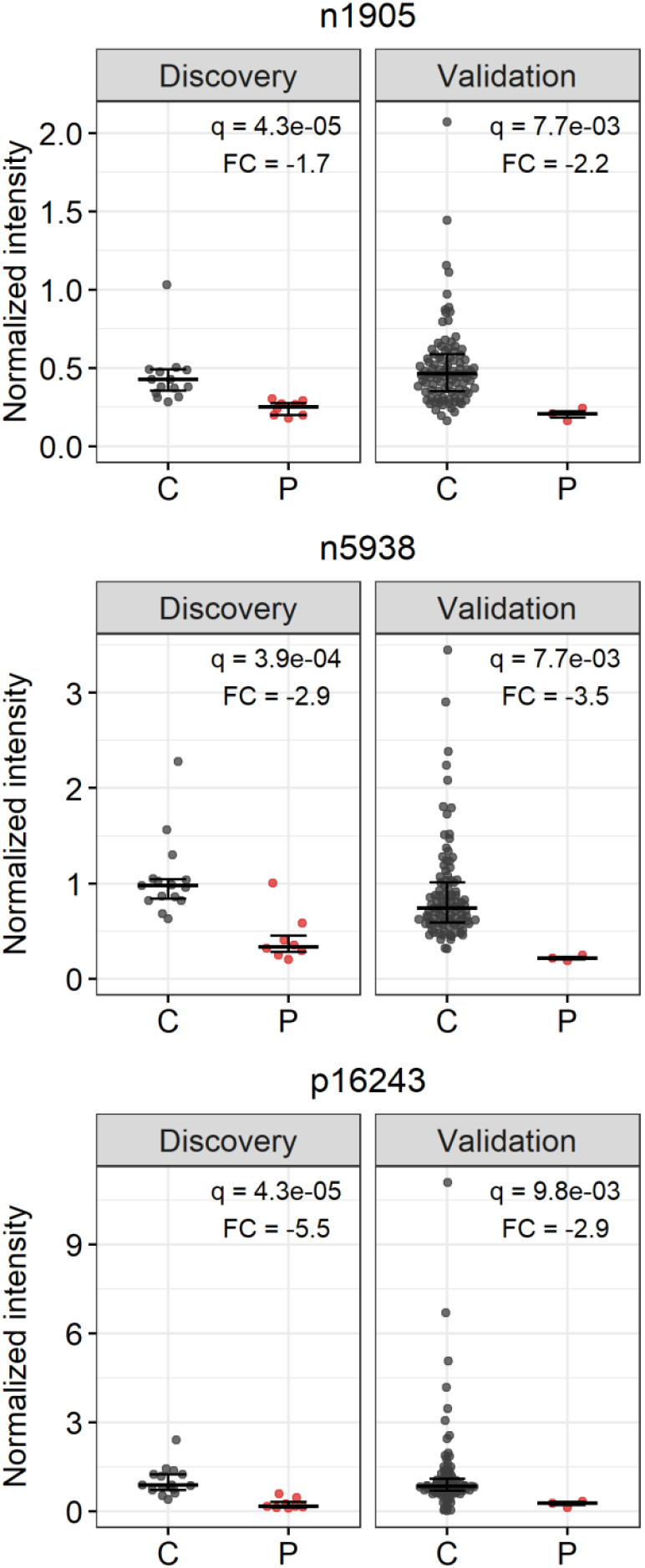
The three selected features n1905, n5938 and p16243 and their normalized intensity in controls (C) and patients (P). Both the ‘discovery cohort’ and ‘validation cohort’ are shown, using QC-based normalization to allow comparing data from different measurements. Bars represent median with interquartile range. For n1905, four outliers in the control cohort, with normalized intensities of 8, 12, 88, and 135 are not shown in the graph, but are included in the statistics. *q* = adjusted *P*-value (Wilcoxon signed-rank test with Benjamini-Hochberg correction), FC = fold change (calculated by median (P) / median (C)).

### Annotation of selected features n1905 (*m/z* 195.0507, RT 0.67)

The m/z resulted in multiple hits in the HMDB: d-mannonic acid, 2-carboxyarabinitol, gulonic acid, (l-)gluconic acid, and galactonic acid (all as [M-H]-adducts). Chemical standards of gluconic and galactonic acid showed a matching RT. Therefore, we analysed six CSF samples (four controls, two patients) using IP-LC-MS/MS which allowed separation of gluconic and galactonic acid. Gluconic acid showed a higher intensity than galactonic acid, but both could be detected. While the intensities of both acids significantly correlated with the intensities of feature n1905 across the six samples individually, the sum of the intensities showed the strongest correlation (r = 0.98, p = 4.3e-4; Fig. 3A+B). This indicates that n1905 is a mixture of these two metabolites, with the major part of the signal representing gluconic acid.

**Figure 3.**
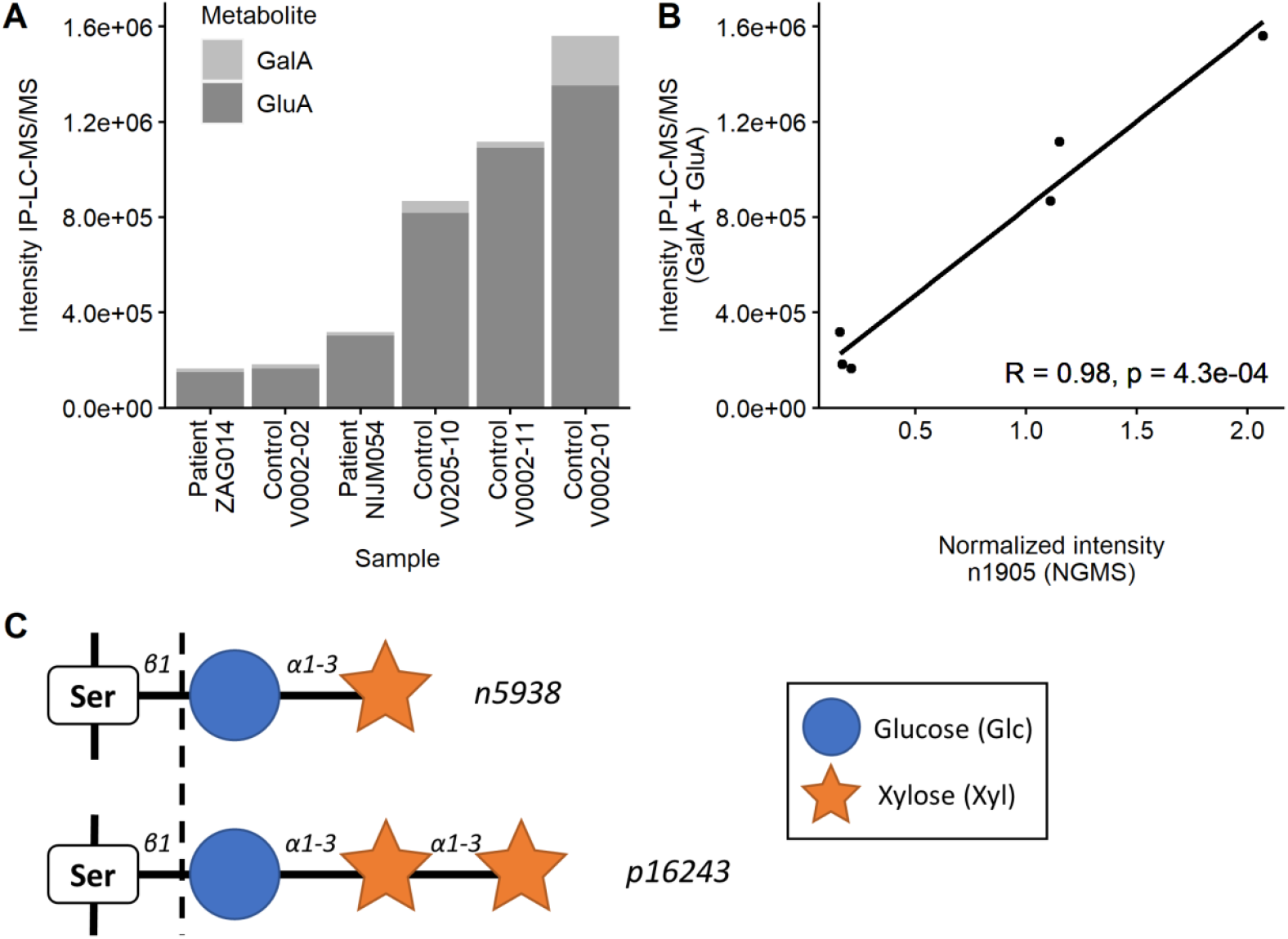
Annotation of selected features. (**A**) Intensities of galactonic acid (GalA) and gluconic acid (GluA) in a subset of six samples (two patients, four controls) as measured by ion-pair liquid chromatography tandem mass spectrometry (IP-LC-MS/MS). (**B**) Correlations between the normalized intensity of n1905 as measured by NGMS and cumulative intensities of GalA and GluA measured by IP-LC-MS/MS. R = Pearson’s correlation coefficient. (**C**) Features n5938 and p16243 were annotated as xylose-α1-3-glucose and xylose-α1-3-xylose-α1-3-glucose, respectively. These oligosaccharides are usually linked to serine (Ser) residues in epidermal growth factor-like repeats of proteins through a process called *O*-glucosylation.^18,19^ However, we found them in an unbound form (indicated by the dotted line).

### n5938 (*m/z* 347.0756, RT 0.81)

Based on the isotope pattern in the raw data mass spectrum, this feature was identified as an [M+Cl]^-^ adduct. A corresponding [M+Na]^+^ adduct was found in the raw data. A HMDB search returned seven metabolites with chemical formula C11H20O10, all representing disaccharides consisting of a combination of a hexose and a pentose. Since the intensity was decreased in GLUT1DS, we hypothesized that the hexose was glucose. Of all known human sugars in glycoconjugates, xylose is the only pentose, present in Xyl-Glc on EGF repeats (Fig. 3C).^18,19^Both Xyl-α1-3-Glc and Xyl-β1-3-Glc were synthesized and compared with n5938 (see Supplementary Material). Exact mass and HILIC-MS/MS confirmed the disaccharide structure. IRIS data then assigned n5938 to the α-stereoisomer, finalizing the identification.

### p16243 (*m/z* 467.1370, RT 1.01)

Feature p16243 was identified as an [M+Na]^+^ adduct, with the corresponding [M+Cl]^-^ adduct being among the significant features as well. Searching the HMDB provided no fitting annotations. However, the most likely predicted formula in MassHunter was C16H28O14, which could fit with a trisaccharide consisting of one hexose and two pentoses. In line with the identified disaccharide, a Xyl-Xyl-Glc trisaccharide, described in human and other species (Fig. 3C), was considered a likely match.^18,19^ A synthesized standard of Xyl-Xyl-Glc showed a match based on exact *m/z*, HILIC RT and IRIS (see Supplementary Material), confirming this annotation of p16243.

### Diagnostic value of selected features

In each patient at least one of the three selected features had a normalized intensity below p5 of the control group. For gluconic + galactonic acid, five patients had values even below p2.5. For Xyl-Glc, all nine values below p5 were also below p2.5. The normalized intensities for Xyl-Xyl-Glc were below p5 in 7 patients, but none were below p2.5. Table 1 includes the results of the three features in all individual patients.

## Discussion

Targeted analysis of NGMS data showed decreased CSF glucose and lactate as known for GLUT1DS, confirming the validity of NGMS to search for GLUT1DS biomarkers. Interestingly, we also demonstrated increased CSF glutamine, previously reported in a single patient with GLUT1DS.^12^ Our data show that while CSF glutamine values may be within the control range for individual patients, they are significantly increased at group level. We have no clear explanation for this observation; the origin and fate of CSF glutamine are determined by multiple metabolic pathways and cannot easily be linked to GLUT1DS.^20^ Using untargeted analysis, we identified decreased levels of CSF gluconic + galactonic acid, Xyl-Glc, and Xyl-Xyl-Glc in GLUT1DS.

Although gluconic acid is commonly encountered in plants and fruits, its metabolic origin in humans is largely unknown.^21^ After phosphorylation, gluconic acid can serve as a substrate for the pentose-phosphate pathway (PPP).^21,22^ Galactonic acid is found in healthy individuals in small amounts, and can also fuel the PPP.^23^ Possibly, in the case of GLUT1DS, the lack of glucose-6-phosphate, the common substrate for the PPP, could be compensated for by alternative substrates like gluconic and galactonic acid.^22^ This mechanism would explain the low CSF concentrations of both metabolites in GLUT1DS. Interestingly, CSF lactate concentrations are thought to be low in GLUT1DS for a similar reason, namely as a consequence of the capability of the brain to use lactate as an alternative fuel for glucose.^24^

The decreased CSF Xyl-Glc and Xyl-Xyl-Glc levels in GLUT1DS may be linked to another fundamental aspect of cell metabolism: *O*-glucosylation, a posttranslational modification at serine and threonine residues of proteins. Xyl-Glc and Xyl-Xyl-Glc glycans are attached to specific sequences in evolutionarily conserved epidermal growth factor-like (EGF) repeats (Fig. 3C), found in secreted proteins and extracellular domains of transmembrane proteins. Among those are Notch receptors, which are known to play a key role in embryonic neurogenesis and cell proliferation, among many other vital cellular processes.^18,19,25-27^ It has been shown that Xyl-Xyl-Glc stabilizes individual EGF repeats, thereby regulating Notch trafficking in cells.^28^ Lack of these specific *O*-glycans may thus disrupt the Notch signalling pathway. Since dozens of other proteins are also *O*-glucosylated, deficient *O*-glucosylation may affect multiple cellular processes apart from Notch signalling.^18,19,25-27^ Additionally, a glucose shortage in the brain may impact the biosynthesis of complex lipid species, such as gangliosides.^29^

While the attachment of Xyl-Glc and Xyl-Xyl-Glc as a posttranslational modification of many human proteins is firmly established, their presence in unbound form in body fluids has not been documented before. Possibly, both oligosaccharides result from the degradation of their proteins of origin. We hypothesize that their low CSF concentrations in GLUT1DS reflect compromised *O*-glucosylation capacity of cells within the CNS, as a consequence of insufficient glucose availability due to defective transport into the CNS. Although the decrease in CSF xylose concentration in GLUT1DS did not reach significance (Fig. 1), GLUT1 deficiency may contribute to diminished xylose transport and if so, brain *O*-glucosylation would be compromised by limited availability of both glucose and xylose.

A limitation of this study was the relatively small number of CSF samples, necessitating future studies in larger patient cohorts to determine the exact diagnostic values of (the combination of) various CSF biomarkers in GLUT1DS. Also, a larger control sample size should be evaluated for possible age-dependency of biomarker concentrations, the influence of KDT, adequate CSF fraction to be collected, and possible effects of fasting before lumbar puncture. Furthermore, our samples had variable preanalytical conditions. While this did not directly negatively influence our study, a prospective design and standardized sample collection and storage methods are preferred for future studies.

In conclusion, our NGMS methodology confirmed the typical diagnostic CSF profile of GLUT1DS with low values of glucose and lactate and increased glutamine. Importantly, we identified three novel CSF biomarkers of GLUT1DS: gluconic + galactonic acid, Xyl-Glc, and Xyl-Xyl-Glc. All were significantly decreased in CSF of patients compared to controls. Our findings improve the diagnostic accuracy of CSF analysis in GLUT1DS, and provide new leads towards a full understanding of the multiple roles of glucose in the brain and how these different roles may be compromised in GLUT1DS. As such, this study guides therapy development and adds new biomarkers for monitoring of novel therapeutic strategies.

## Abbreviations

2DOG: 2-deoxy-D-glucose
3OMG: 3-O-methyl-D-glucose
DHA: dehydroascorbic acid
DKG: diketogulonic acid
EGF: epidermal growth factor-like
GLUT1DS: glucose transporter type 1 deficiency syndrome
HILIC-IRIS: hydrophilic interaction liquid chromatography infrared ion spectroscopy
HMDB: human metabolome database
IEM: inborn error of metabolism
IP-LC-MS/MS: ion-pair liquid chromatography tandem mass spectrometry
KDT: ketogenic diet therapy
*m/z*: mass-to-charge ratio
NGMS: next-generation metabolic screening
PPP: pentose-phosphate pathway
QC: quality control
RPLC-MS: reversed-phase liquid chromatography mass spectrometry
RT: retention time
Xyl-Glc: xylose-α1-3-glucose
Xyl-Xyl-Glc: xylose-α1-3-xylose-α1-3-glucose

## Acknowledgements

The authors would like to thank Anahita Rassi (Division of Clinical Chemistry and Biochemistry of the University Children’s Hospital Zürich) for her help in sample selection and collection of clinical data.

## Funding

The study was funded by Stofwisselkracht under the project name of “Innovative diagnostics in cerebrospinal fluid of patients with neurometabolic disorders” (KLM Coene and MM Verbeek). MAAP Willemsen received an unrestricted grant from the Glut1 Deficiency Foundation (Owingsville, USA). Furthermore, the authors gratefully acknowledge the Nederlandse Organisatie voor Wetenschappelijk Onderzoek (NWO) for the support of the FELIX Laboratory. This work was supported by the NWO division Exact and Natural Sciences (grant numbers TTW 15769, TKI-LIFT 731.017.419) and Radboud University through an interfaculty collaboration grant. The authors confirm independence from the sponsors; the content of the article has not been influenced by the sponsors.

## Competing interests

The authors report no competing interests.

## Supplementary material

Supplementary material is available online.

## Notes

### Competing Interest Statement

The authors have declared no competing interest.

### Author Declarations

The Commissie Mensgebonden Onderzoek Radboudumc of the Radboud University Medical Center gave ethical approval for this work.

