## Supplementary Material for "Novel CSF biomarkers of GLUT1 deficiency syndrome: implications beyond the brain’s energy deficit"

#### Extended methods

##### Synthesis of DKG

To synthesize diketogulonic acid (DKG), an ice-cold solution of (L)-dehydroascorbic acid was hydrolysed with KOH, precipitated form cold ethanol and quickly filtered as reported, considering the inherent instability of the product (Figure S1).[1]

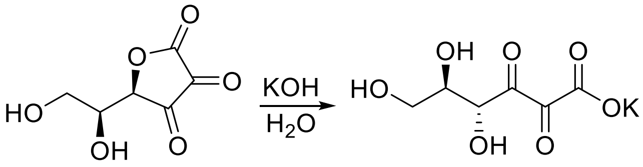


*Figure S1. Synthesis of diketogluconic acid (right) from (L)-dehydroascorbic acid (left).*

##### Synthesis of (Xyl-α1-3-)Xyl-α1-3-Glc

The full synthesis is described in reference 2. In short, glycosylation of perbenzylated Xylose thioglycoside donor with commercially available 1,2:5,6-di-O-isopropylidene-α-D-glucofuranose acceptor resulted, after deprotection, in the isolation of both α and β-linked disaccharides Xyl-α1-3-Glc and Xyl-β1-3-Glc. For the trisaccharide an initial glycosylation of the 1,2:5,6-di-O-isopropylidene-α-D-glucofuranose acceptor with a 3' orthogonal protected Xylose thioglycoside donor resulted in the intermediate disaccharide. Orthogonal deprotection of the 3' position of this disaccharide and further glycosylation of this acceptor with the perbenzylated Xylose thioglycoside donor yielded the desired protected trisaccharide. Final deprotection resulted in the isolation of Xyl-α1-3-Xyl-α1-3-Glc.

##### Analysis of hexonic acids by IP-LC-MS/MS

CSF samples were prepared as for UHPLC-QTOF-MS, but dissolved in pure Milli-Q instead of 0.1% v/v HCOOH in Milli-Q at the last step. The samples were analyzed by targeted ion-pair liquid chromatography tandem mass spectrometry (IP-LC-MS/MS) using the Agilent 1290 Infinity UPLC and 6490A Triple Quadrupole Mass Spectrometer (QQQ-MS) (Agilent, Santa Clara, CA, USA). Five microliters of sample was injected into a HSS T3 column (100 Å, 1.8 µm, 2.1 mm X 150 mm; Waters, Milford, Massachusetts, USA) and hexonic acids were separated isocratically using 10 mM tributylamine, 12 mM acetic acid, 2 mM acetylacetone and 3% MeOH as the mobile phase. QQQ-MS operated in multiple-reaction monitoring (MRM) mode using 195.1->129.0 and 195.1->75.0 transitions for selective detection of hexonic acids. Intensities of galactonic and gluconic acid were extracted using Skyline 20.2 and were subsequently correlated to UHPLC-QTOF-MS intensities using Pearson's correlation coefficient.

##### Hydrophilic interaction liquid chromatography infrared ion spectroscopy (HILIC-IRIS)

All samples were separated via a hydrophilic interaction liquid chromatography (HILIC) method for improved separation and detection, combined with infrared ion spectroscopy (IRIS). For HILIC-IRIS analysis, the CSF samples were dissolved in Milli-Q, identical to the analyses by ion-pair LC-MS/MS described above. From there, acetonitrile and ammonium hydroxide was added to a final composition of 80% acetonitrile (v/v) and 0.1% ammonium hydroxide. All synthesized standards were dissolved in 80:20 acetonitrile:water with 0.1%

ammonium hydroxide. The mobile phase gradient went from 80% acetonitrile with 0.1% ammonium hydroxide to 30% acetonitrile with 0.1% ammonium hydroxide in 7 minutes at a flowrate of 200 µl/min, followed by 16 minutes of re-equilibration. These conditions result in a significant proportion of ammoniated species in the ESI-MS, which was recently found to be favorable for IRIS identification of sugar molecules.[2]

Between 2 and 20 µl of sample was injected per analysis on a Waters BEH Amide column (100 x 2.1 mm i.d., 1.7 µm particles) by use of a UHPLC system (Bruker Elute). A loop-switching method was used to generate signal from one HILIC separated feature for long enough to record an IRIS spectrum. The IRIS experiments were performed on a modified quadrupole ion trap mass spectrometer (Bruker, amaZon Speed ETD) with optical access to the trapped ion population.[3,4] In the ion trapping region, mass-to-charge selected ions were irradiated by the wavelength-tunable infrared free electron laser FELIX to induce wavelength-dependent photodissociation. IRIS spectra were obtained by plotting the photo-dissociation yield as a function of IR wavelength and were linearly corrected for the wavelength-dependent infrared laser pulse energy.[3,5]

### Extended results

#### Hydrophilic interaction liquid chromatography infrared ion spectroscopy (HILIC-IRIS)

MS/MS showed exact matches between the n5938 and p16243 features and their respective synthesized standards. MS/MS could not distinguish between the alpha and beta disaccharide isomer. Confirmed MS/MS fragments are listed in Table S1.

*Table S1: Measured* m/z *values for feature precursor ions and tentatively assigned fragments*

| Analyte | Precursor ion (PI) | Characteristic fragments |
|---|---|---|
| n5938, Xyl-β1-3-Glc, Xyl-α1-3-Glc | 335.095 $[M+Na]^+$ | 317.084 (PI-$H_2O$); 203.05 ($[hexose+Na]^+$); 185.041 ($[hexose-H_2O+Na]^+$); |
| | 330.140 $[M+NH_4]^+$ | 295.015 (PI-$H_2O$-$NH_3$); 163.012 ($[hexose-H_2O+H]^+$); 115.013 ($[pentose-(H_2O)_2+H]^+$); |
| p16243, Xyl-α1-3-Xyl-α1-3-Glc | 467.154 $[M+Na]^+$ | 449.143 (PI-$H_2O$); 335.105 (PI-(pentose-$H_2O$)); 287.082 (PI-hexose); 203.058 $[hexose+Na]^+$; |
| | 462.199 $[M+NH_4]^+$ | 445.171 (PI-$NH_3$); 427.159 (PI-$H_2O$-$NH_3$); 295.111 (PI-pentose-$H_2O$-$NH_3$); |

The HILIC retention times (RTs) of n5938 and p16243 matched their synthesized standards (7.54 and 4.51 min, respectively), except for Xyl-β1-3-Glc which had a slightly different RT (7.26 min), not fully resolved from n5938. The concentration of feature n5938 was estimated at <10 nM in patient control CSF. The IRIS spectrum of feature n5938 can be found in Figure S2. The black trace represents a single measurement on a patient sample, whereas the blue and green traces represent the averages of three measurements per synthesized compound. A frequency range of approximately 300-1000 $cm^{-1}$ was chosen for IRIS analysis, because this range was established to be highly diagnostic for saccharide-like molecules.[2] Figure S2 shows a clear match between the Xyl-α1-3-Glc synthesized compound (in green) and n5938, whereas the Xyl-β1-3-Glc deviates most notably in the 700-950 $cm^{-1}$ range. Based on these results, we confidently assign the Xyl-α1-3-Glc molecular structure to feature n5938.

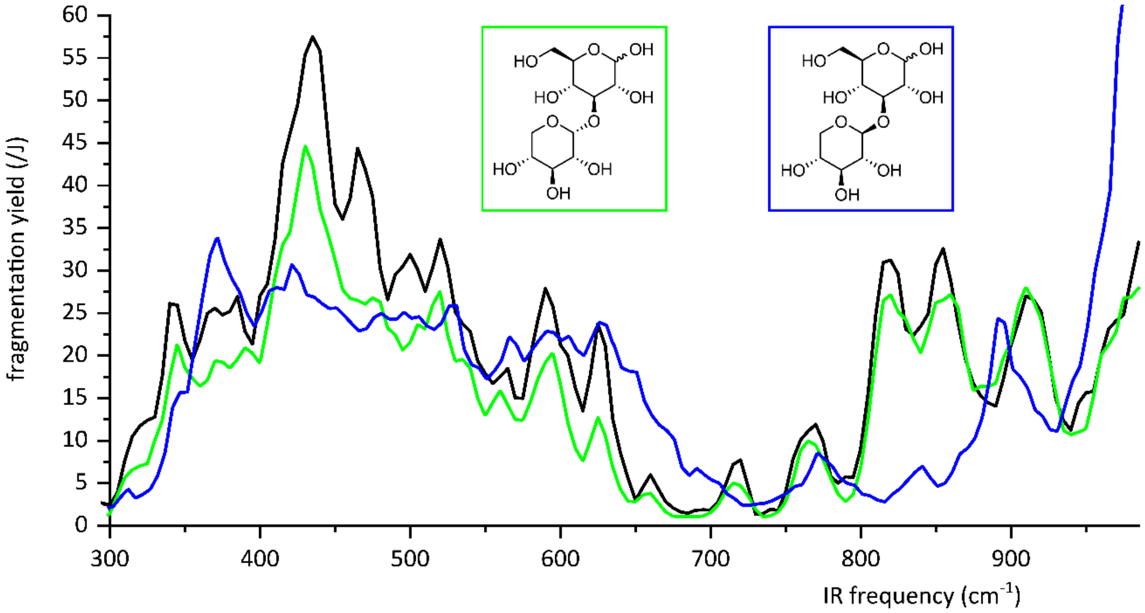


*Figure S2. Isomeric identification of feature n5938 by comparison of IRIS data with synthetic reference standards. The experimental IR spectra of the [M+NH4]+ species (m/z 330.1395) of feature n5938 (black trace) and synthetic reference compounds (Xyl-α1-3-Glc and Xyl-β1-3-Glc, green and blue traces, respectively) are shown. Structural formulae of the reference compounds are inlaid.*

Figure S3 shows the experimental spectra of the Xyl-α1-3-Xyl-α1-3-Glc synthesized compound (in purple) and feature p16243 (in black), the average of 4 and 6 measured spectra, respectively. We applied a correction to compensate for the presence of an isobaric instrument interference due to the low abundance of feature p16243 (estimated at < 5 nM). A more elaborate discussion of the IRIS method and results can be found in reference 2.

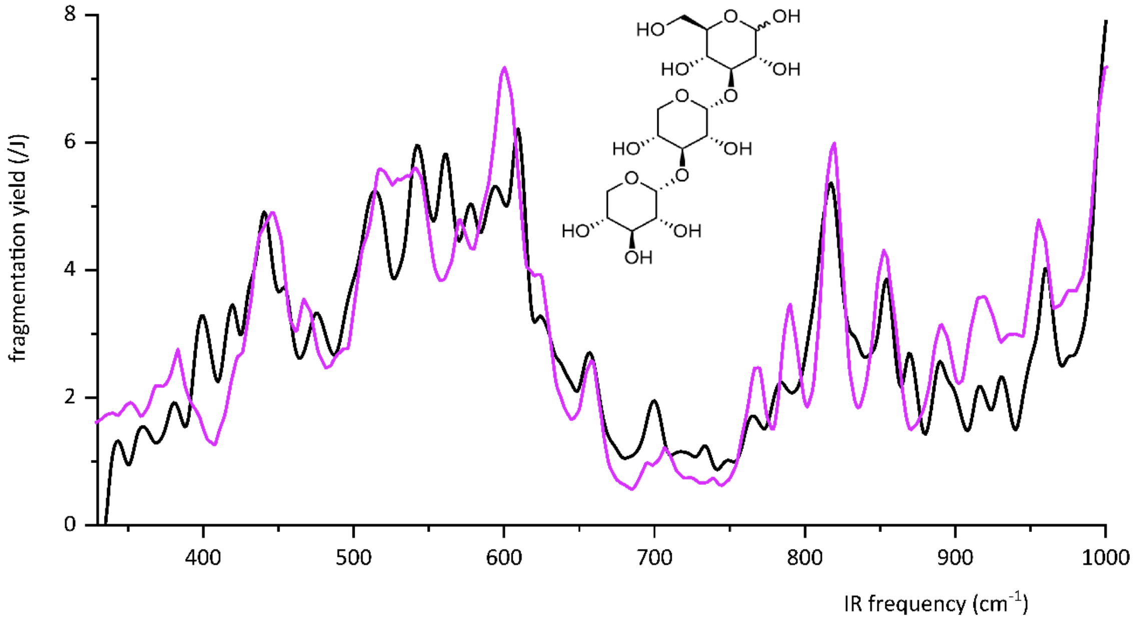


*Figure S3. Isomeric identification of feature p16243 by comparison of IRIS data with synthetic reference standards. The experimental IR spectra of the [M+NH4]+ species (m/z 462.1370) of feature p16243 (black trace) and Xyl-α1-3-Xyl-α1-3-Glc reference compound (purple trace) are shown. Structural formula of the assignment is inlaid.*
